# Towards the integrative theory of Alzheimer’s disease: linking molecular mechanisms of neurotoxicity, beta-amyloid biomarkers, and the diagnosis

**DOI:** 10.1101/2022.12.07.22283236

**Authors:** Yaroslav I. Molkov, Maria V. Zaretskaia, Dmitry V. Zaretsky, the Alzheimer’s Disease Neuroimaging Initiative

**Author notes:** Data used in the preparation of this article were obtained from the Alzheimer’s Disease Neuroimaging Initiative (ADNI) database (adni.loni.usc.edu). As such, the investigators within the ADNI contributed to the design and implementation of ADNI and/or provided data but did not participate in analysis or writing of this report. A complete listing of ADNI investigators can be found at: http://adni.loni.usc.edu/wp-content/uploads/how_to_apply/ADNI_Acknowledgement_List.pdf.

## Abstract

A major gap in amyloid-centric theories of Alzheimer’s disease (AD) is that even though amyloid fibrils *per se* are not toxic *in vitro*, the diagnosis of AD clearly correlates with the density of beta-amyloid (Aβ) deposits. Based on our proposed amyloid degradation toxicity hypothesis, we developed a mathematical model explaining this discrepancy. It suggests that cytotoxicity depends on the cellular uptake of soluble Aβ rather than on the presence of amyloid aggregates. The dynamics of soluble Aβ in the cerebrospinal fluid (CSF) and the density of Aβ deposits is described using a system of differential equations. In the model, cytotoxic damage is proportional to the cellular uptake of Aβ, while the probability of an AD diagnosis is defined by the Aβ cytotoxicity accumulated over the duration of the disease. After uptake, Aβ is concentrated intralysosomally, promoting the formation of fibrillation seeds inside cells. These seeds cannot be digested and are either accumulated intracellularly or exocytosed. Aβ starts aggregating on the extracellular seeds and, therefore, its concentration decreases in the interstitial fluid. The dependence of both Aβ toxicity and aggregation on the same process – cellular uptake of Aβ – explains the correlation between AD diagnosis and the density of amyloid aggregates in the brain.

We tested the model using clinical data obtained from the Alzheimer’s Disease Neuroimaging Initiative (ADNI), which included records of beta-amyloid concentration in the cerebrospinal fluid (CSF-Aβ42) and the density of beta-amyloid deposits measured using positron emission tomography (PET). The model predicts the probability of AD diagnosis as a function of CSF-Aβ42 and PET and fits the experimental data at the 95% confidence level.

Our study shows that existing clinical data allows for the inference of kinetic parameters describing beta-amyloid turnover and disease progression. Each combination of CSF-Aβ42 and PET values can be used to calculate the individual’s cellular uptake rate, the effective disease duration, and the accumulated toxicity. We show that natural limitations on these parameters explain the characteristic distribution of the clinical dataset for these two biomarkers in the population.

The resulting mathematical model interprets the positive correlation between the density of Aβ deposits and the probability of an AD diagnosis without assuming any cytotoxicity of the aggregated beta-amyloid. Finally, to the best of our knowledge, this model is the first to mechanistically explain the negative correlation between the concentration of Aβ42 in the CSF and the probability of an AD diagnosis.

## INTRODUCTION

Dr. Alzheimer first described extracellular senile plaques and intracellular neurofibrillary tangles as specific to a certain kind of dementia [3] later named Alzheimer’s Disease (AD) [25]. The main component of the extracellular deposits is beta-amyloid protein (Aβ) [23, 24]. Aβ was found to be toxic to cultured cells [14, 17, 38]. Also, histopathological changes are more likely to occur in cells in close proximity to the plaques [10, 37, 60]. Based on this evidence, the etiology and pathogenesis of AD were associated by the scientific community with the Aβ plaques. Further, by means of the positron emission tomography (PET) technique, the density of amyloid deposits was measured in live subjects, which brought more indirect data supporting a potential role of Aβ in AD pathogenesis [42] as cognitively impaired AD patients had significantly denser amyloid deposits. Even more importantly, patients with a significant density of amyloid deposits appeared much more prone to decline cognitively in the future compared to amyloid-negative individuals [50].

The density of amyloid deposits negatively correlates with Aβ42 concentration in the cerebrospinal fluid (CSF) [21, 41, 64]. Due to the strength of this negative correlation, the overall diagnostic accuracy of CSF-Aβ42 and PET studies is considered similar [42]. However, CSF-Aβ42 levels and PET imaging data on the density of amyloid deposits were recently shown to have independent predictive powers for different Alzheimer’s disease markers, including AD diagnosis itself, hippocampal volume, and brain metabolism [41]. Importantly, patients with AD tend to have lower CSF-Aβ42 levels than research subjects with normal cognition, even if their PET levels are the same [41, 59].

The decrease of CSF-Aβ42 with an increased density of amyloid deposits can be explained by a more intense aggregation of soluble amyloid on already aggregated amyloid. However, the aggregation cannot explain why decreased CSF-Aβ42 is associated with a higher probability of AD diagnosis at the same levels of amyloid load and hence the same aggregation rate. There are other factors that can affect the Aβ concentration in the CSF such as the Aβ synthesis rate and the Aβ filtering rate largely defined by the CSF flow intensity. Aβ synthesis is not lower in patients with AD [43], and the CSF flow is not more intense in patients with AD [22, 58]. Therefore, a decreased CSF-Aβ42 can only be explained by more intense clearance of the peptide inside the brain [72].

The most obvious mechanism of intratissue removal of Aβ42 is cellular uptake. The link between increased cellular uptake of beta-amyloid and increased neuronal death can be explained by the amyloid degradation toxicity hypothesis [70]. After endocytosis, beta-amyloid is digested by lysosomal proteases. Some peptide fragments can aggregate into oligomeric forms capable of creating membrane channels and thus permeabilizing lysosomal membranes which happen to have all the properties required to incorporate amyloid membrane channels [68, 69]. Lysosomal permeabilization is a well-established consequence of the exposure of cells to beta-amyloid [34, 67]. Amyloid channels can be extremely large [6, 39, 45], which would allow, according to our estimates, for the release of lysosomal cathepsins into the cytoplasm [71]. Cytoplasmatic leakage of lysosomal proteases can activate necrotic and/or apoptotic processes [26, 32, 36, 61], and thus be the molecular mechanism initiating neuronal death. However, amyloid degradation toxicity hypothesis *per se* does not explain why the density of amyloid deposits correlates with the probability of AD diagnosis or has any prognostic value.

To better understand the nature of the correlation between the density of amyloid deposits, Aβ42 concentration in the CSF, and the progression of AD, we developed an integrative mathematical model describing beta-amyloid homeostatic regulation and its connection to the diagnosis of AD. Based on the above, we assumed that cellular amyloid uptake is an important factor defining Aβ toxicity *in vivo*. The cognitive effect of Aβ toxicity accumulates over time, so cognitive decline is faster when cellular amyloid uptake is higher. The degree of neuronal damage induced by Aβ uptake varies between individuals due to either differences in the sensitivity to toxic action of beta-amyloid or various mechanisms of protective response. Importantly, we hypothesized that due to the intensity of protein turnover, aggregation seeds do not form in the interstitial fluid, but appear first intracellularly from the endocytosed Aβ. After being exocytosed, these seeds grow by aggregating soluble Aβ and become senile plaques. This hypothesis implicates cellular uptake as an underlying mechanism for both Aβ aggregation and cytotoxicity. In the present study, we used these assumptions to map the inferred beta-amyloid kinetics to the probability of AD diagnosis and compared the output of the model to the existing clinical dataset containing data on the Aβ concentration in the CSF and the density of amyloid deposits.

## METHODS

### Clinical dataset

We used non-personalized data available through the Alzheimer’s Disease Neuroimaging Initiative (ADNI) (http://adni.loni.usc.edu/). The ADNI was launched in 2003 as a public-private partnership, led by Principal Investigator Michael W. Weiner, MD. The primary goal of ADNI has been to test whether serial magnetic resonance imaging, positron emission tomography (PET), other biological markers, and clinical and neuropsychological assessment can be combined to measure the progression of mild cognitive impairment (MCI) and Alzheimer’s disease (AD). The study protocol for ADNI was approved by the local ethical committees of all participating institutions and all participants signed informed consent, which included consent for de-identified data being shared with the general scientific community for research purposes [2]. The authors of this manuscript did not participate in data acquisition and received de-identified data after approval by ADNI.

Our analysis included all ADNI participants for whom the ascertainment of normal cognition (NC) or AD diagnosis, as well as CSF collection, were made within one year from a PET scan identifying brain amyloidosis. The number of research subjects in the AD and NC groups was 143 and 476, respectively. All subjects were evaluated between June 2010 and February 2019. Amyloid positivity was defined by PET data according to ADNI guidelines as a standard uptake value ratio (SUVR) at or above 1.08 for [18]Fflorbetaben or 1.11 for [18]F-florbetapir, with a higher SUVR indicating a greater amyloid plaque burden. Details regarding PET acquisition are described in previous publications and on the ADNI website (www.adni-info.org). Given the use of two different amyloid PET-tracers, SUVR levels were converted to centiloids (CL) using the specific equation for each tracer as provided by ADNI. Datapoints included in this study are shown in Fig. 1.

**Figure 1.**
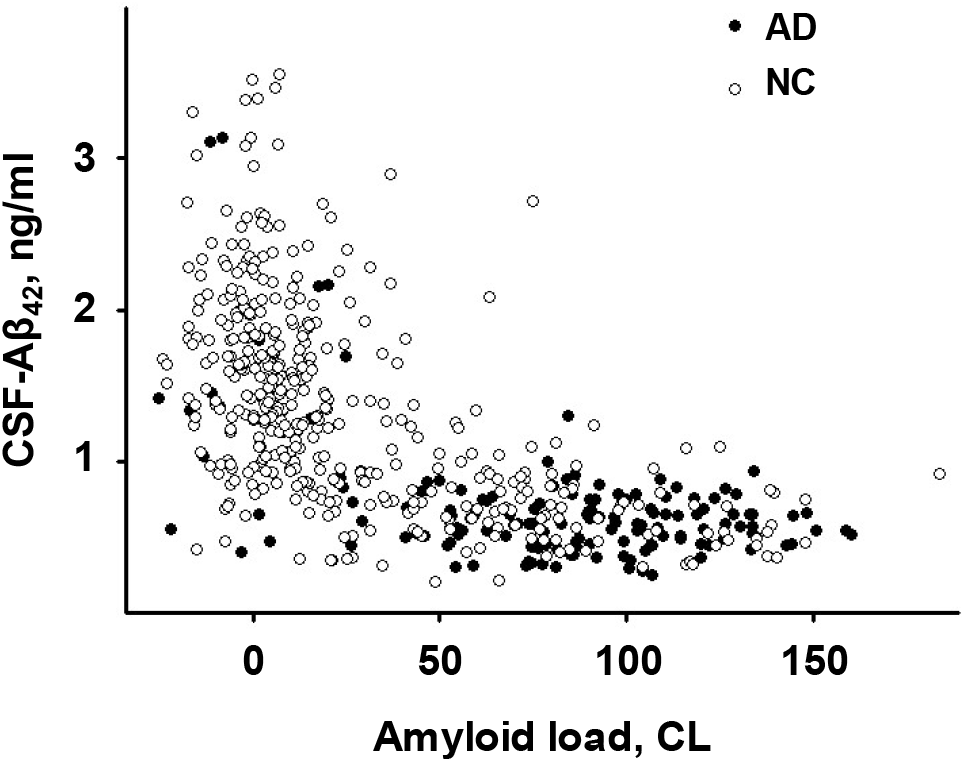
ADNI data on CSF-Aβ42 vs beta-amyloid load in research subjects with normal cognition (NC, open circles) and patients with Alzheimer’s disease (AD, closed circles).

### A mathematical model of the cerebral amyloid turnover

The model employed in this study extends the ordinary differential equation (ODE) model we used previously [72]. The concentration of soluble Aβ in the interstitial fluid (ISF), denoted by [*ISF*], is controlled by several processes: (1) synthesis by cells, (2) filtration of the protein into the CSF, (3) aggregation into non-soluble plaques, and (4) uptake by cells (Fig. 2). Beta-amyloid taken up by cells either is digested and initiates cytotoxicity or aggregates into fibrils which can be exocytosed. Cytotoxic insult is proportional to the amount of endocytosed CSF-Aβ42. In turn, exocytosed fibrils serve as seeds for beta-amyloid aggregation from the interstitial fluid. The rate of removal of soluble Aβ from the solution increases with the growth of the deposits.

**Figure 2.**
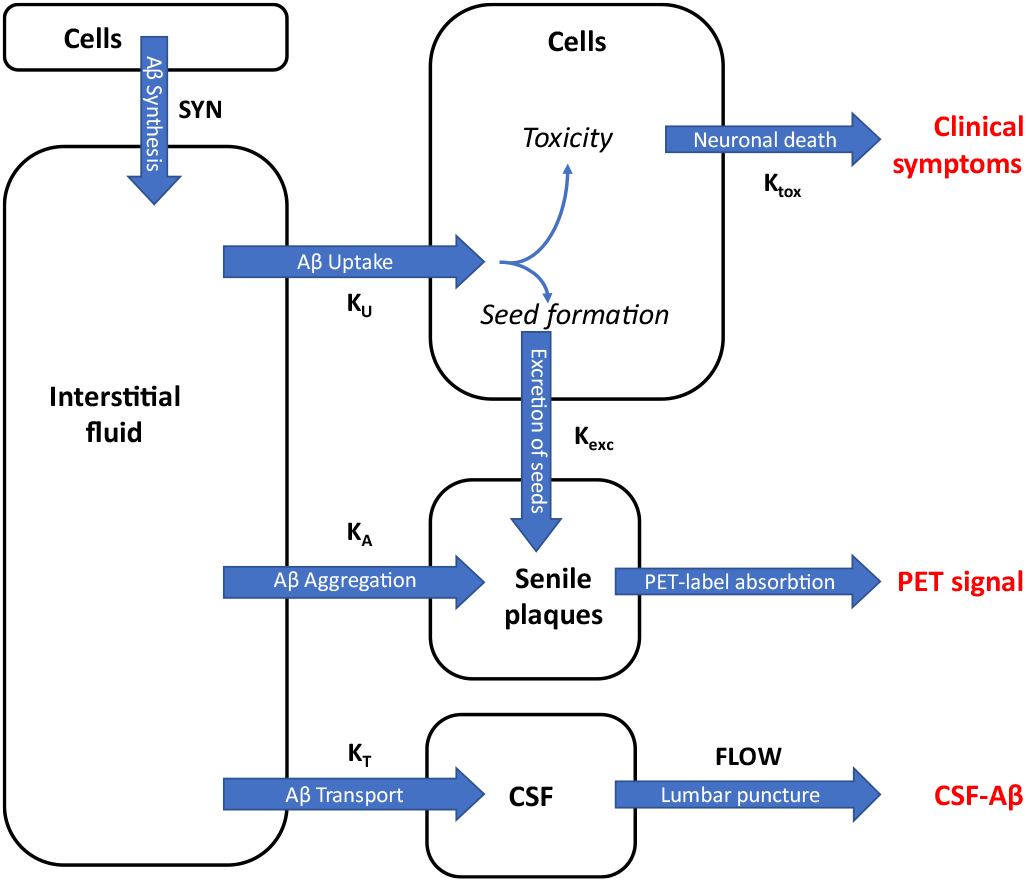
Schematic of the model of beta-amyloid turnover describing the mathematical relationship between Aβ cytotoxicity, which leads to Alzheimer’s disease (AD), and two biomarkers of AD: CSF-Aβ42 (measured in the CSF collected by lumbar puncture) and amyloid load in the brain (measured using positron-emission tomography). The parameters of the model are shown next to the arrows.

The model and its analysis are based on the following assumptions:

1. Synthesis rate (*SYN*) is independent of both interstitial Aβ42 and the density of plaques.
2. The rate of removal of protein through the CSF is a product of the CSF flow (*FLOW*_*CSF*_) and CSF-Aβ42 (*CSF*): *FLOW*_*CSF*_ · [*CSF*].
3. The model assumes a linear relationship between the concentrations of soluble Aβ42 in the ISF and the CSF with a coefficient of transfer *K*_*T*_: *CSF*= *K*_*T*_· [*ISF*].
4. The rate of cellular uptake of soluble Aβ42 (*uptake*) is proportional to the interstitial Aβ42 concentration [*ISF*] with a coefficient of uptake *K*_*u*_ : *K*_*u*_ · [*ISF*].
5. The exocytosis of intracellularly produced fibrils is proportional to the *uptake*, and therefore is proportional to [*ISF*]: *K*_*e*_· *uptake* = *K*_*e*_· *K*_*u*_ · [*ISF*].
6. Existing plaques serve as seeds for the aggregation of soluble Aβ42 in the ISF. The rate of loss of soluble Aβ42 from the ISF due to aggregation is a product of Aβ42 concentration in the ISF, the concentration of plaques ([*PET*], calculated from the intensity of the PET signal), and the coefficient of aggregation *K*_*a*_ : *K*_*a*_ · [*PET*] · [*ISF*].
7. Current intensity of toxic insult (current toxicity) is proportional to the uptake: *K*_*tox*_ · *uptake*.
8. Neural damage (accumulated toxicity, *TOX*) is an integral of current toxicity over the duration of illness (*T*_*ill*_).

Under these assumptions, the system of ordinary differential equations describing the dynamics of beta-amyloid concentrations in the ISF and CSF, and the density of amyloid deposits characterized by the PET signal will have the following form:

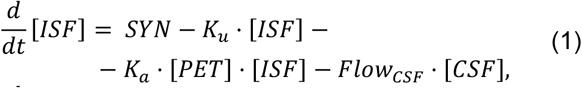

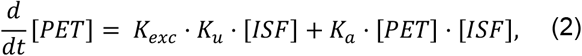

where *CSF*= *K*_*T*_· [*ISF*], with an initial condition [*PET*] = 0 at *t* = 0. Assuming that the time scale of soluble Aβ42 dynamics is much shorter than the time scale of beta-amyloid deposits accumulation, so at each time Aβ42 concentration in ISF/CSF is at instantaneous equilibrium, one can show that *CSF*and [*PET*] satisfy the following relationship:

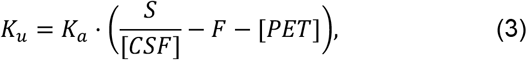

where *F* and *S* are compound parameters proportional to *Flow*_*CSF*_ and *SYN*, respectively: *F* = *K*_*T*_/*Ka* · *Flow*_*CSF*_ ; *S* = *K*_*T*_/*Ka* · *SYN*. Using the above relationship, Eq. (2) can be solved, and explicit expressions for the accumulated toxicity and the disease duration can be obtained:

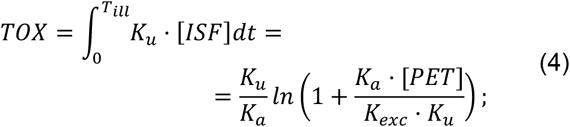

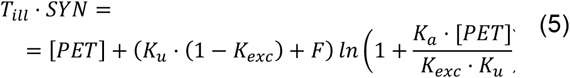

Note, Eqs. (3) and (4) together allow calculating the accumulated toxicity for an individual based on their current readings of the CSF-Aβ42 and Aβ deposit density (the intensity of PET signal).

### Mapping the accumulated toxicity to the probability of AD diagnosis

We assume that each individual has their own toxicity threshold *T*, so they become sick with AD if their accumulated toxicity exceeds this threshold. We further assume that the probability distribution of the toxicity thresholds in the population is normal with a certain mean *T*_0_ and variance 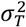. Therefore, the probability of AD based on the individual’s accumulated toxicity can be calculated using the cumulative distribution function (CDF) of the normal distribution as follows:

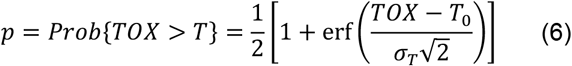

where erf(.) is the error function.

### Parameter inference

In terms of the number of participants, the ADNI dataset we used is biased towards patients with AD diagnosis (∼26%) compared to 10% of similar age patients in the general population [1]. To account for this discrepancy, we created a surrogate dataset by triplicating records of the healthy participants. This resulted in the corrected fraction of AD patients in the group reflecting the one observed in the general population.

For each participant, the AD diagnosis *T*is a Boolean random variable having the Bernoulli distribution (a particular case of the binomial distribution *B*(1, *p*)) with the above probability, which is a function of Aβ42 concentration in CSF, the PET signal as well as 5 unknown parameters, *T*_0_, *σ*_*T*_, *Kexc, S* and *F*. AD diagnoses of different participants are assumed independent. Therefore, the likelihood function for the ADNI dataset is as follows:

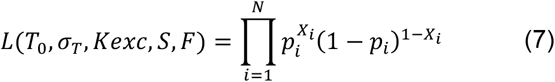

where *N* is the total number of datapoints, *X*_*i*_ = 1 if the *i*^*th*^ participant is diagnosed with AD, *X*_*i*_ = 0 otherwise, and *p*_*i*_ = *p*([*PET*]_*i*_, [*CSF*]_*i*_ ; *T*_0_, *σ*_*T*_, *Kexc, S, F*) is the probability of AD diagnosis given the measured values [*PET*]_*i*_ and [*CSF*]_*i*_ of the *i*^*th*^ participant as defined by Eq. (6). To identify the parameter values, we used the Maximum Likelihood Estimation (MLE) method. Hereinafter we refer to those as the MLE parameter values. In this procedure, datapoints which either had negative PET values or resulted in negative values of the uptake rate were assumed outliers and were excluded from the consideration.

### Model’s goodness-of-fit evaluation

To evaluate how well the model output corresponds to the ADNI data, we used Pearson’s chi-square test. We partitioned the ([*PET*], [*CSF*])-plane into non-overlapping rectangular cells with dimensions of 25 CL by 500 pg/ml. We calculated the number of AD patients (*x*_*i*_) and cognitively normal participants in each cell. These numbers were corrected by the factors of 0.4 and 1.2, respectively, to adjust the fraction of AD patients to 10% (see above), while keeping the overall number of participants the same (see the supplemental spreadsheet for calculations). We used Eq. (6) to calculate the expected number of AD patients in the center of each cell as a product of the total number of participants in this cell (*n*_*i*_) and the probability of AD diagnosis (*p*_*i*_), as predicted by the model with MLE parameter values. Then we constructed a chi-square statistic as follows:

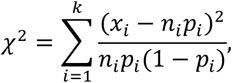

where *k* = 23 is the number of cells with non-zero number of participants. If the model fits well, the calculated statistic should be a sample from a chisquare distribution. The number of degrees of freedom of the chi-square distribution equals the number of cells used (23 cells) minus the number of model parameters subject to optimization (5 parameters). Using this distribution, we calculated the p-value as the probability of the chi-square statistic to exceed the calculated value. We accepted 0.05 as the significance level.

## RESULTS

### The model accurately predicts the fraction of AD patients based on the values of PET and CSF-Aβ42

We found the following values of the model parameters using the Maximum Likelihood Estimation approach (see Methods): *S* = 250 CL·ng/ml, *F* = 78 CL, *K*_*e*_= 0.47, *T*_0_ = 190 CL, and *σ*_*T*_= 87 CL.

Figure 3A shows the probability of an AD diagnosis, calculated using these parameter values, as a function of CSF-Aβ42 and amyloid load (PET). The region of possible PET and CSF-Aβ42 values has an upper boundary representing a zero-uptake rate (*K*_*u*_ = 0), i. e. *S*/*CSF* − *F* − *PET* = 0. Experimental data are superimposed onto the theoretical probability distribution with measurements obtained from participants with normal cognition shown in green, and measurements obtained from AD patients shown in yellow. Several experimental points that appear above the theoretical upper boundary are marked as outliers and removed from consideration. Goodness-of-fit of the model was evaluated using Pearson’s chi-square test (*p* = 0.19, see Methods, Supplemental Materials). We conclude that the model prediction of AD diagnosis based on the PET and CSF-Aβ42 readings fits the experimental data at a 95% confidence level.

**Figure 3.**
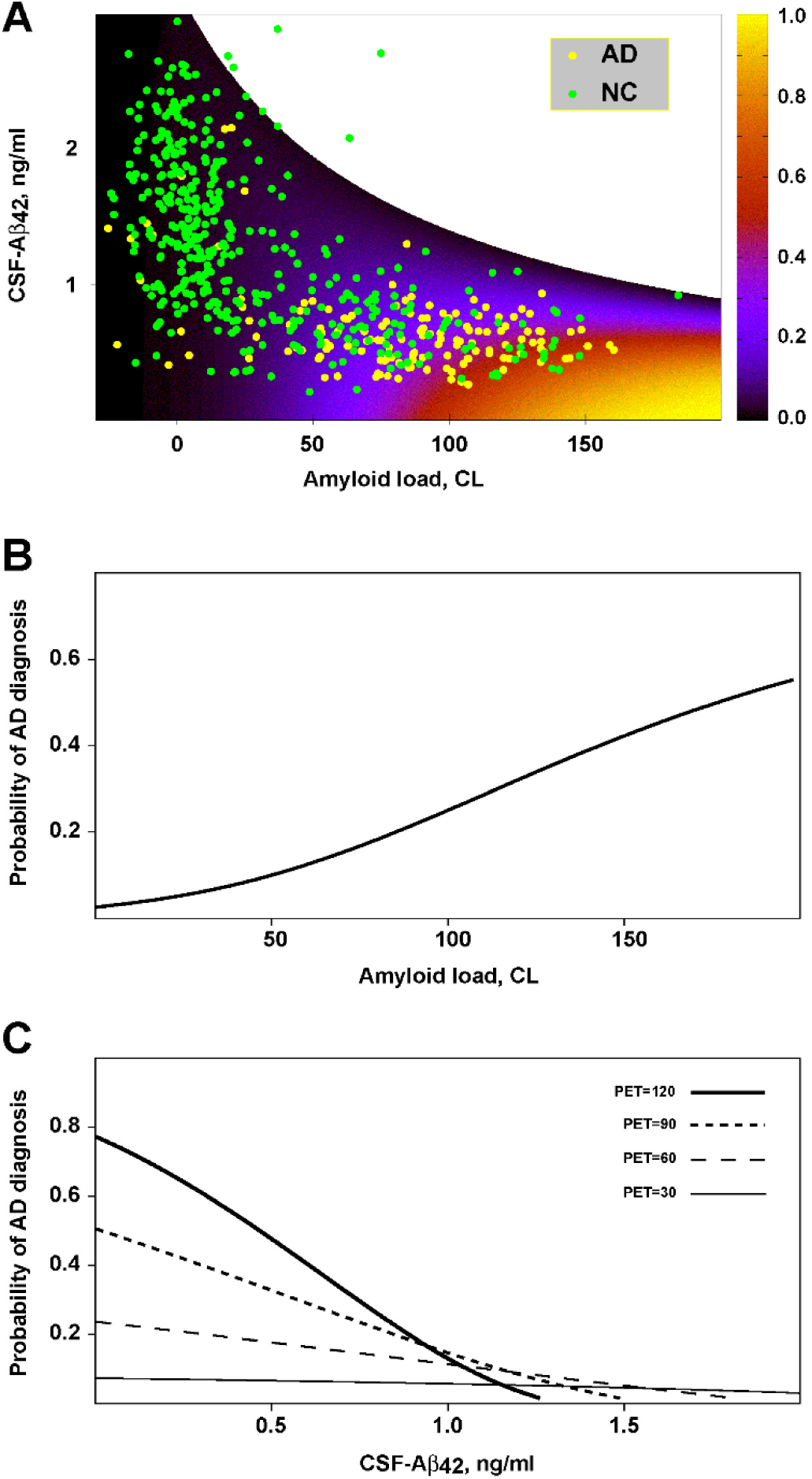
The model tuned to clinical data. **A**. The probability of Alzheimer’s disease as a function of CSF-Aβ42 and amyloid load based on the best-fit parameters of the model. The overlain scatter plot shows data points indicating subjects with normal cognition (NC, green circles) and patients with Alzheimer’s disease (AD, yellow circles). **B**. Probability of AD diagnosis as a function of betaamyloid load calculated using the model under the assumption that, for any given amyloid load, the values of CSF-Aβ42 are uniformly distributed in the range corresponding to the experimentally measured values. **C**. Probability of AD diagnosis as a function of CSF-Aβ42 for various levels of amyloid load.

### Positive correlation between the AD diagnosis and the density of amyloid deposits

The proposed model readily explains the positive correlation between an AD diagnosis and the density of amyloid deposits without a causal effect of amyloid deposits on neurotoxicity. Indeed, if CSF-Aβ42 levels are not considered, the probability of AD diagnosis, as calculated by the model, increases with amyloid load (Fig. 3B).

The total amyloid uptake can be represented as a product of the time-averaged cellular uptake rate and the duration of disease. Since our model allows for estimating the latter (see Eq. (3) in Methods), we analyzed curves representing the hypothetical AD “age” of participants (Fig. 3B) calculated based on their current PET and CSF-Aβ42 readings. Unsurprisingly, longer disease durations generally correspond to greater accumulated amyloid deposits. However, this accumulation depends on CSF-Aβ42 in a non-trivial way (see Eq. (2) in Methods). Specifically, for long-enough disease durations (see curve 3.5 in Fig. 4B), the accumulated deposits are maximal at intermediate CSF-Aβ42 values (0.5-1 ng/ml). In general, CSF-Aβ42 negatively correlates with the cellular amyloid uptake rate (see more on that below), so an increase in CSF-Aβ42 corresponds to an effective decrease in cellular amyloid uptake, and, therefore, a slower excretion of amyloid seeds (see Fig. 2). On the other hand, extremely low CSF-Aβ42 levels lead to a reduction in amyloid aggregation on already excreted seeds and somewhat lower PET levels for the same disease duration (see the lower part of the curve 3.5 in Fig. 4B).

**Figure 4.**
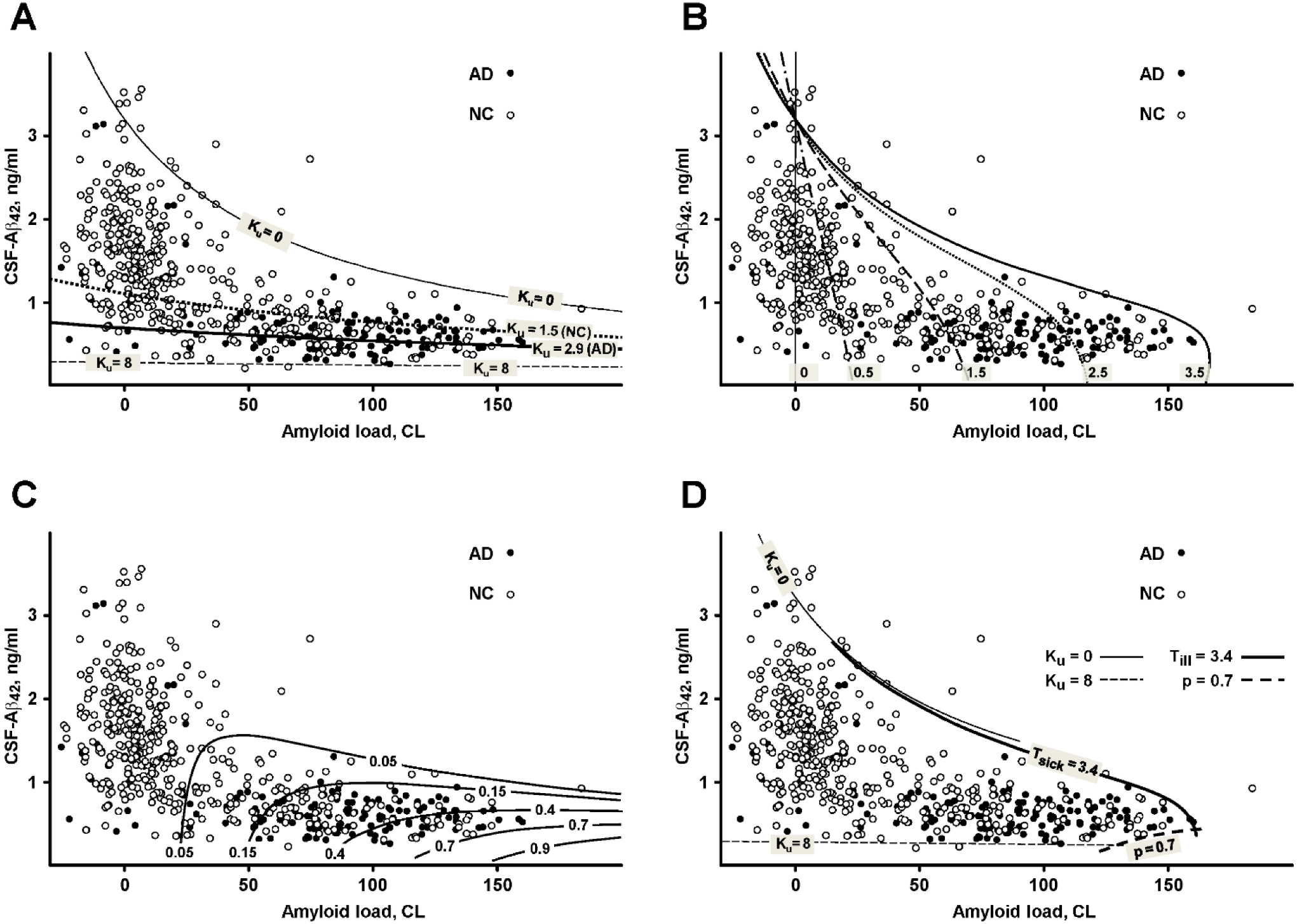
Level curves for various values of the cellular uptake rate, the disease duration and the diagnosis probability in relation to the distribution of clinical data points. The uptake rate and the disease duration are in arbitrary units. The distributions of datapoints from research subjects with normal cognition (NC, open circles) and patients with Alzheimer’s disease (AD, closed circles) are shown as the scatter plot of CSF-Aβ42 (ng/ml) vs beta-amyloid load (centiloids, CL). **A**. Level curves for the uptake rate are ordered from top to the bottom with an increasing value of the uptake rate. The curves corresponding to the average uptake rates for NC subjects and AD patients are shown as dotted and thick solid lines, respectively. **B**. Level curves of the disease duration indicated by the labels next to the curves. **C**. Level curves of the probability of AD diagnosis labelled by the corresponding probability values. They alsorepresent the accumulated toxicity level curves with the corresponding accumulated toxicity values. **D**. The cloud of datapoints in the clinical dataset is bounded by (1) minimal and maximal cellular uptake rate values (solid and dashed thin lines, correspondingly); 2) the maximal disease duration (thick solid line labeled T_ill_= 3.4); (3) highest accumulated toxicity (thick dashed line; the probability of AD diagnosis p = 0.7).

### Negative correlation between CSF-Aβ42 and AD diagnosis

Another important clinical observation that can be explained by the model is that, at the given density of amyloid deposits, the probability of AD diagnosis increases as CSF-Aβ42 reduces (Fig. 3C). According to the model, at the highest possible levels of CSF-Aβ42, the probability of AD diagnosis is low due to a small cellular uptake rate. An increase in amyloid removal through uptake leads to lower CSF-Aβ42, as well as to elevated neurotoxicity. Again, our model assumes that lower CSF-Aβ42 levels *per se* are not causal to more intensive neurodegeneration. Rather, both processes result from the increased cellular uptake.

Level curves corresponding to several different uptake rate values (see Eq. (1) in Methods) are shown in Fig. 4A. At the uppermost curve labeled “K_u_ = 0” the uptake rate equals 0, and all data points appearing above this curve are considered outliers (see Methods) since, according to the model, values of CSF-Aβ42 above this curve would correspond to negative values of the uptake rate. As we increase the amyloid uptake rate, the corresponding level curves occur progressively lower, reflecting a reduction in CSF-Aβ42 caused by higher uptake. At the same time, the probability of AD diagnosis increases (Figs. 3A,C) due to the increased cellular uptake of amyloid. This explains why the average uptake rate in the AD group appears higher than in the participants with normal cognition [72] (Fig. 4A). We illustrated this phenomenon by showing the level curves corresponding to the uptake rate averages for cognitively normal participants and AD patients (K_u_ = 1.5 and K_u_ = 2.9, respectively, in Fig. 4A). At extremely high uptake values, the level curves become nearly horizontal and serve as a lower boundary for CSF-Aβ42 (see K_u_ = 8 in Fig. 4A).

### The model explains the shape of the dataset on the (PET, CSF-Aβ42)-plane

The distribution of data points on the (PET, CSF-Aβ42)plane has a characteristic shape (Figs. 1, 4). Natural limitations on the parameters of the model can serve as the boundaries of this cloud (Fig. 4D). As noted, the upper boundary results from the fact that the cellular uptake rate cannot be negative. Therefore, physiologically plausible values of CSF-Aβ42 must satisfy the inequality

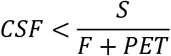

which follows from Eq. (1) in Methods (see K_u_ = 0 in Fig. 4A).

The lower boundary in terms of CSF-Aβ42 appears horizontal, i.e., the lowest possible values of CSF-Aβ42 are almost independent of the density of amyloid deposits characterized by PET. This lower boundary corresponds to highest possible uptake rate values that make intratissue clearance much greater than the aggregation (see K_u_ = 8 in Fig. 4A). Highest uptake corresponds to an approximate minimal possible CSF-Aβ42 value of 0.3 ng/ml (which is approximately equal to *S*/8).

On the right, the cloud is bounded by the maximum possible PET values, which correspond to the largest possible density of amyloid deposits. Accumulation of the aggregated amyloid is limited by the disease’s lifespan. Therefore, the right boundary can be described by the level curve of the disease duration corresponding to the participants with the longest times of disease progression. Note that the curve corresponding to the longest disease progression also serves as an upper boundary (compare curves T_ill_ = 3.4 and K_u_ = 0 in Fig. 4D).

Finally, it is reasonable to expect that very high accumulated toxicity levels should be incompatible with survival, and, therefore, we would not see participants corresponding to those levels. Figure 4C shows the level curves for accumulated toxicity/AD diagnosis probability. One can observe that indeed, the curve corresponding to an AD diagnosis probability of 0.7 fits well as the lower boundary of the cloud around 150 CL PET values (see curve p = 0.7 in Fig. 4C).

## DISCUSSION

Recent history of the scientific progress in Alzheimer’s disease research is filled with controversies regarding the role of the most prominent and well-known anatomical correlate of the disease, the amyloid plaques. Even though the density of the plaques has a strong correlation with the clinical status of the patient, not all patients with a high amyloid load have the disease [20]. Also, the Aβ concentration used in cell cultures for the toxicity studies exceeds the naturally observed one by several orders of magnitude (ng/ml in the CSF [4, 21, 59] vs μg/ml in *in vitro* toxicity experiments [17, 38, 56]). The lack of/minimal effectiveness of the drugs targeting Aβ in clinical trials [12, 19, 66] further undermines the confidence that beta-amyloid deposits are in fact relevant to AD etiology and pathogenesis.

Nevertheless, the correlations between the presence of Aβ aggregates in the brain, the concentration of Aβ42 in the CSF, and the clinical status of the patients are obvious and beg for some mechanistic explanations. Considering that Aβ is, in fact, toxic to cells [14, 17, 38], we have built a mathematical model connecting molecular mechanisms of amyloid toxicity to clinical data contained in the ADNI database available to researchers through an extensive public-private partnership [46, 47]. Here, we discuss the basis of our model’s assumptions and how it resolves the controversies.

### Main assumptions of the model

#### Mechanism of beta-amyloid toxicity

The cytotoxic effect of Aβ in the model is based on the amyloid degradation toxicity hypothesis [70]. This hypothesis has the following cornerstones.

First, the molecular mechanism of Aβ toxicity is the permeabilization of cellular membranes by membrane channels formed by amyloid oligomers. This is similar to the membrane channel hypothesis first introduced in the early 1990s by Arispe et al. [5-7, 52].

Second, we assume that channels are formed by Aβ degradation products. We previously demonstrated that the 42 amino acids-long Aβ peptide does not effectively form membrane channels, while the undecapeptide Aβ25-35 does [68, 69]. The latter is one of the products of the proteolytic degradation of the full-length peptide, also known for its extreme neurotoxicity [44]. So, unlike the original amyloid membrane channel hypothesis, we assume that the fragments (such as Aβ25-35), rather than the full-length peptide, are largely responsible for the formation of the channels. It is possible that membrane channels can be formed by various fragments [33, 35, 45].

Third, the channels occur in the lysosomal membranes, rather than in the plasma membrane. Due to a positive charge of the Aβ25-35, these fragments can form channels in the negatively charged membranes only [29, 68, 69]. The outer leaflet of plasma membrane is mostly neutral in healthy cells [8]; however, some intracellular membranes, such as the inner mitochondrial and lysosomal membranes, have a significant negative surface charge [68, 70]. While the delivery route to the inner mitochondrial membranes is unclear, the lysosomal membranes are obvious targets for Aβ-induced damage. Indeed, Aβ was previously shown to be endocytosed and found intralysosomally [34, 65, 67]. Next, lysosomes contain various proteases and are responsible for peptide degradation with Aβ25-35 being one of the products [44]. It is quite possible that some other Aβ fragments can also form membrane channels [35].

These three basic assumptions can qualitatively explain several important features of AD, such as lysosomal dysfunction, mitochondrial damage, increased apoptosis, etc. [16, 18, 27, 49, 54, 55]. They also imply that the neurodegeneration rate depends on the intensity of the lysosomal permeabilization process, which is in turn defined by the Aβ influx through endocytosis. In line with that, in the model, the toxic effect of Aβ is proportional to the amount of endocytosed peptide (see Fig. 2). Throughout the manuscript, we refer to this process as cellular amyloid uptake.

#### Formation of amyloid deposits

Aβ is synthesized in the form of propeptide that undergoes proteolytic processing and is released into the interstitial fluid [28]. In terms of concentration, a major form of beta-amyloid is Aβ40 (40 amino acids-long peptide). However, unlike Aβ40, a second most prominent form, Aβ42 (42 amino acids-long peptide), has a strong correlation with AD diagnosis [48, 57]. Aβ42 is also more prone to aggregation through the formation of beta-sheets [48].

The process of Aβ aggregation is non-linear. Aβ is synthesized and released into the interstitial fluid without any specific conformation. To initiate the aggregation process, it is necessary to create aggregation seeds first, which grow quickly after that. In experimental conditions, at the concentration of peptide in a μM range, the seeding of Aβ42 takes many hours [15, 31]. By extrapolating to the interstitial concentration of beta-amyloid *in vivo*, which is in an nM range, the seeding would take several months [15]. This makes spontaneous seeding in the interstitial space negligible over the typical time interval between the appearance of the monomeric molecule in the ISF and its clearance from the ISF through any mechanism: the half-life of brain Aβ42 is measured in hours [51]. However, after endocytosis, the intralysosomal concentration of amyloid is almost 100 times higher than its extracellular concentration [30, 65]. In lysosomes, the peptide remains enclosed intravesicularly for more than 24 hours [13, 65], thus providing the proper conditions and time needed for seed formation. The resistance of aggregated Aβ42 to proteolysis explains its intracellular accumulation [30]. Non-digestible inclusions are exocytosed; in the case of amyloid aggregates, they serve as aggregation seeds in the extracellular space attracting and appending the soluble Aβ [11].

Thus, in the model, the rate of change of the amyloid deposit density has two terms (see Eq. (2) in Methods, Fig. 2). The first one represents the intracellular formation of amyloid fibrils, whose intensity is proportional to the cellular amyloid uptake. The second term represents the aggregation process on the already-formed deposits, with a rate proportional to the product of the interstitial Aβ42 concentration and the current amyloid deposit density.

#### Translating neurotoxicity into AD diagnosis

As noted, in the model, the intensity of the toxic insult is proportional to the rate of amyloid uptake by neurons, and the ultimate neuronal damage accumulates over time. However, the total amount of Aβ consumed by neurons may not define the degree of neuronal death by itself due to individual differences in sensitivity (resistivity) to its toxicity for the following reasons. First, the concentration of the toxic amyloid forms may depend on the balance between their creation and degradation. The fragmentation of the full-length peptide is mediated by exoproteases, while further degradation of fragments is performed by endoproteases. These processes may be mediated by different enzymes (even though some lysosomal proteases have both endo- and exo-proteolytic activity). The balance of these two proteolytic activities may have individual-to-individual variability. Also, any biochemical damage caused may be offset by protective and/or reparatory processes. For example, the damage from leaking cathepsins can be prevented by cytoplasmic protease inhibitors, such as cystatins [40]. Most likely, there are many other factors affecting the relationship between the intensity of Aβ uptake and neurotoxicity.

To account for these individual factors in the model, the development of the disease is considered probabilistic. Specifically, the overall toxic effect is proportional to the amyloid uptake accumulated over time (see Eq. (4) in Methods). The individual threshold for the accumulated toxicity leading to an AD diagnosis is assumed to be randomly distributed over the population. This way, the probability for a participant with a given accumulated toxicity to have an AD is numerically equal to the cumulative distribution function of the distribution of the thresholds (see Eq. (6) in Methods).

### The model explains previously paradoxical clinical observations related to the density of amyloid deposits and Aβ42 concentration in the CSF

As noted, one of the greatest controversies in the field is that the occurrence of Alzheimer’s disease and its progression strongly correlates with the accumulation of extracellular amyloid deposits [42, 50], despite the aggregated Aβ is not toxic *in vitro* by itself [17, 38]. Our model accurately reproduces this correlation (Figs. 3A, B) without any underlying assumptions about the neurotoxicity of the amyloid deposits. In the model, both amyloid aggregation and its toxicity depend on the internalization of soluble Aβ. Therefore, an increase in the cellular amyloid uptake intensifies the accumulation of both amyloid deposits and its neurotoxic effects (Fig. 2). Both elevated neurodegeneration and higher accumulation of amyloid plaques in AD originates from increased cellular amyloid uptake. The fact that neuronal death and the accumulation of amyloid plaques have the same origin manifests as a positive correlation between the probability of AD diagnosis and amyloid load (Fig. 3B).

Another strikingly paradoxical observation is that Alzheimer’s disease and its progression are associated with the decreased concentration of soluble Aβ42 in the cerebrospinal fluid [21, 41, 64], despite, unlike aggregated Aβ, soluble Aβ forms are cytotoxic i*n vitro* [14, 17, 38]. Due to aggregation of soluble Aβ42 on the exocytosed seeds and existing plaques, the concentration of soluble Aβ42 in the interstitial liquid becomes lower at higher amyloid load levels. Considering that the density of plaques positively correlates with the probability of AD diagnosis, it is not too surprising that CSF-Aβ42 levels are lower in AD patients. However, CSF-Aβ42 is lower in AD patients compared to subjects with normal cognition, even if they have the same amyloid load [41, 59]. This seems counterintuitive, as lower CSF-Aβ42 implies lesser Aβ42 availability for endocytosis. However, our model reproduces a negative correlation between CSF-Aβ42 and AD probability at a fixed amyloid load too (Fig. 3C). The reason for this phenomenon is that lower CSF-Aβ42 levels are in this case caused by higher cellular amyloid uptake rates, resulting in an overall greater amount of amyloid being endocytosed and, therefore, its increased toxicity.

### The model describes the distribution of PET and CSF-Aβ42

The distribution of clinical data in coordinates (PET, CSF-Aβ42) is not uniform (Fig. 1). Specifically, CSF-Aβ42 has a wide range of values at low levels of PET. With increased PET, the CSF-Aβ42 range becomes progressively smaller due to decreasing maximal CSF-Aβ42 values, whereas the lower CSF-Aβ42 boundary remains relatively constant. Our analysis of the model shows that the upper and lower boundaries of this data cloud match the curves corresponding to a zero and a largest possible value of the cellular amyloid uptake rate, respectively (Fig. 4A). The zero-uptake curve has a hyperbolic shape reflecting a reduction in the maximal physiologically possible values of CSF-Aβ42 with an increasing amyloid load, while the maximum uptake line (dashed line in Fig. 4A) is almost horizontal. The right boundary of the cloud in Fig. 1 corresponding to the maximal PET values fits well with the longest disease progression time. We considered a family of isochrones on the (PET, CSF-Aβ42)-plane corresponding to progressively longer sickness (Fig. 4B) and noticed that one of them can serve as the right boundary of the cloud (Fig. 4C). Based on this, we can speculate that the right boundary corresponds to the longest times of the disease’s progression, which may correlate with the maximal age of the participants. Note, however, that we do not make any assumptions in the model about the link between the length of the disease and the biological age.

### The model suggests new AD biomarkers

In this study, we developed a theoretical basis for correlations between AD diagnosis probability and two extensively used AD biomarkers, Aβ42 concentration in the CSF and the density of amyloid deposits characterized by PET. Using the model, we identified two independent factors defining this probability that are the individual rate of the cellular amyloid uptake and disease duration. The latter is not equivalent to the patient’s biological age as the amyloid accumulation is known to start in middle age [9]. In the model, we can interpret the initiation of the disease as a stepwise increase in the cellular amyloid uptake rate at some point in life, which jumpstarts the disease progression. Therefore, monitoring and/or controlling this parameter may be instrumental in AD screening/ managing at very early stages. While there are no ready-to-use techniques to measure the intrabrain Aβ42 clearance, the SILK technique could be a good starting point [53].

In the model, we describe the AD diagnosis probabilistically, meaning that the same accumulated toxicity may or may not result in AD in a given individual. This implies that people have different toxicity thresholds quite broadly distributed over the population. Therefore, AD prevention can be based on heightening the amyloid toxicity thresholds. For example, per the amyloid degradation toxicity hypothesis, cytotoxicity occurs when specific amyloid fragments form membrane channels in the lysosomal membranes [70]. Correspondingly, the balance of endo-/exoproteolytic activities promoting or preventing the formation of channel-forming fragments could be a predictor of AD progression rate. These channels allow for the leakage of lysosomal cathepsins, which in turn activates the cell’s necrosis or apoptosis. So, one of the ways to fight these toxic effects is to neutralize leaked cathepsins using some protease inhibitors such as cystatins [62].

Clearly, exploiting any of these routes requires new extensive studies focused on amyloid endocytosis and the mechanisms of amyloid cytotoxicity.

### The model predicts no positive outcome from dissolving amyloid deposits

Since AD was associated with amyloid plaques, amyloid deposits have been pharmacological targets in hopes that their dissolvement can slow down disease progression. Our model offers a new perspective on this intensely-debated topic. Considering that amyloid deposits are not toxic in the model, their disappearance will not have any consequences by itself. However, a decrease in the aggregated amyloid concentration will reduce the aggregation rate of soluble amyloid and thus increase its concentration in the ISF and CSF. The general relationship between CSF-Aβ42 and the density of amyloid deposits is provided by Eq. (3). According to this equation, dissolving the existing deposits should increase CSF-Aβ42, which is in fact observed clinically after dissolving senile plaques after the treatment with monoclonal anti-amyloid antibodies [12]. Our model’s prediction is that the dissolvement of the amyloid plaques can only accelerate AD progression. From this perspective, not only is dissolving plaques not helpful but potentially harmful, and a high density of amyloid plaques appears protective rather than dangerous.

Our model does not consider the plaques being toxic by themselves, which is not necessarily the case. For example, it was previously suggested that amyloid deposits could activate inflammatory responses or be the source of oxidative stress due to absorbed metal ions [63]. In these cases, dissolving the deposits would obviously prevent their own toxicity, and the net outcome would depend on the balance between the toxic and protective influences of the deposits in a particular patient.

## Supporting information

Data and statistics

## Data Availability

All data produced in the present work are contained in the manuscript.

https://adni.loni.usc.edu/

## Abbreviations

AD: Alzheimer’s disease
NC: normal cognition
Aβ: beta-amyloid
Aβ42: Aβ1-42
CSF: cerebrospinal fluid
CSF-Aβ42: concentration of Aβ42 in the CSF
PET: positron emission tomography.

## Acknowledgements

Research reported in this publication was not supported by any external funding.

Data collection and sharing for this project was funded by the Alzheimer’s Disease Neuroimaging Initiative (ADNI) (NIH Grant U01 AG024904) and DOD ADNI (award number W81XWH-12-2-0012). ADNI is funded by the National Institute on Aging, the National Institute of Biomedical Imaging and Bioengineering, and through generous contributions from the following: AbbVie; Alzheimer’s Association; Alzheimer’s Drug Discovery Foundation; Araclon Biotech; BioClinica, Inc.; Biogen; Bristol-Myers Squibb Company; CereSpir, Inc.; Cogstate; Eisai Inc.; Elan Pharmaceuticals, Inc.; Eli Lilly and Company; EuroImmun; F. Hoffmann-La Roche Ltd and its affiliated company Genentech, Inc.; Fujirebio; GE Healthcare; IXICO Ltd.; Janssen Alzheimer Immunotherapy Research & Development, LLC.; Johnson & Johnson Pharmaceutical Research & Development LLC.; Lumosity; Lundbeck; Merck & Co., Inc.; Meso Scale Diagnostics, LLC.; NeuroRx Research; Neurotrack Technologies; Novartis Pharmaceuticals Corporation; Pfizer Inc.; Piramal Imaging; Servier; Takeda Pharmaceutical Company; and Transition Therapeutics. The Canadian Institutes of Health Research is providing funds to support ADNI clinical sites in Canada. Private sector contributions are facilitated by the Foundation for the National Institutes of Health (www.fnih.org). The grantee organization is the Northern California Institute for Research and Education, and the study is coordinated by the Alzheimer’s Therapeutic Research Institute at the University of Southern California. ADNI data are disseminated by the Laboratory for Neuro Imaging at the University of Southern California.

The authors thank Daniel Zaretsky for his editorial help.

